# Audio-Visual Stimulation Therapy for Chronic Neuropathic Pain: A Sham-Controlled Randomized Clinical Trial

**DOI:** 10.1101/2024.08.12.24311569

**Authors:** Laura Tabacof, Rebecca Howard, Jeffrey Bower, Erica Breyman, Sophie Dewil, Jenna Tosto-Mancuso, Richard Hanbury, Brandon Carmouche, Mark Robberson, Adam Fry, David Putrino

**Affiliations:** Department of Rehabilitation and Human Performance, Icahn School of Medicine at Mount Sinai, New York, NY; Sana Health, Inc, Louisville, CO

## Abstract

Neuropathic Pain (NP) affects 10% of the general population, decreasing quality of life for millions of Americans and contributing to higher physical and mental health care costs. The most widely used treatments for NP involve medications that show limitations in efficacy and burdensome side effects. This randomized controlled trial explored the efficacy of a wearable Audio-Visual Stimulation neuromodulation device (Sana) as a novel intervention for chronic NP in 64 participants. Outcomes were assessed at baseline, after 8-weeks of daily use of the assigned Sana or Sham device, and after 4 weeks of discontinued use. For the main outcome (Neuropathic Pain Symptom Inventory total), there were statistically significant improvements in the Active arm that were greater than those in the Sham Arm at Week-14 (Mean Difference = 10.04, p = 0.01). Both groups showed significant improvements at the end of the treatment period (Week-10), and the Active arm maintained this improvement after an additional 4 weeks of non-use, while the Sham arm almost returned to baseline (Active Change = 13.26, p <=0.001 | Sham Change = 3.22, p = 0.214). Participants in the Active arm had significant decreases in use of anxiolytic, opiate, antidepressant, and anticonvulsant medications compared to the Sham arm. The study provides strong evidence supporting the efficacy of a novel AVS Device in generating durable improvements in NP, with superiority over Sham at 14 weeks. The Sana device may also reduce the reliance on pain medications and is a safe and easy to use treatment option for patients.

## Introduction

Neuropathic Pain (NP) is defined as a lesion or disease of the somatosensory system. NP affects up to 10% of the general population decreasing the quality of life for millions of Americans while contributing to higher health care costs, estimated to be up to $30,000 in direct and indirect costs annually [2,18,21,27,44,54].

NP presents symptoms such as burning, pressing, tingling or freezing sensations that are often intense enough to disrupt daily activities [3,14,19,52]. Treatments for NP include tricyclic antidepressants, anticonvulsants, and serotonin-norepinephrine reuptake inhibitors which have limited efficacy, burdensome side effects, and drug resistance [3,11,37]. Up to 50% of patients with chronic neuropathic pain are found to be non-adherent to their prescribed treatment regimens [3]. Opiate medications are also commonly prescribed to those with NP, which is effective for reducing pain, but comes with stigma, dependency, and side effects [8,33,46]. The limitations associated with current treatments emphasize the need for innovative and effective treatment strategies for NP.

Audio Visual Stimulation (AVS) is an approach that works by stimulating the central nervous system with patterns of light and sound presented to the eyes patient. The AVS creates a Cortical Evoked Responses (CER) leading to a Frequency Following Responses (FFR) that propagates through the central nervous system. This process induces mental states that are comparable to states of meditation, deep relaxation, rapid sleep onset, reduced anxiety, and reduction of pain [43,47,50]. AVS has shown to induce electroencephalogram (EEG) activity similar to those seen in therapeutic restful states has been associated with reductions in pain (both short- and long-term), depression, anxiety, and insomnia [23,40]. Treatment with AVS may lead to the long-term re-organization of dysfunctional pain processing pathways by affecting the neural circuits responsible for the perception of pain [26,31]. Due to the potential for beneficial neural plastic changes, the application of AVS may lead to long lasting and durable improvements in pain experienced by those with NP. Despite the emerging promise of AVS, there have been few studies exploring its potential efficacy in treating NP.

The goal of this sham-controlled randomized controlled trial is to explore the efficacy of AVS as a novel intervention for NP.

## Methods

### 1. Study design and setting

This was a double-blinded, parallel-arm, sham-controlled, randomized clinical trial investigating the effects of a novel AVS device on symptoms of chronic neuropathic pain. Study procedures were approved by the Mount Sinai’s Program for Protection of Human Subjects 18-2282 and registered on a national clinical trials registry (ClinicalTrials.gov Identifier: NCT04280562 Remote Participation (Within USA) Trial of the AVS Device (named the “Sana Device” developed by Sana Health, Inc, CO, USA). All participants signed an informed consent document prior to enrolment into the study. Sessions were offered at the clinic or via telehealth.

### 2. Randomization and concealment

Individuals were randomly assigned to one of two groups: AVS device or Sham device. Devices were identical in appearance and were randomly distributed to participants. Neither assessor nor participant had access to device classification and code list was kept secured.

### 3. Participants

Participants were recruited from Mount Sinai outpatient clinics, and through social media outreach. Participants could also self-refer through recruitment through ClinicalTrials.gov (Identifier: NCT04280562). In order to be eligible for this study, participants must have met the following criteria: (1) being 18 years of age or older, (2) having a confirmed clinical diagnosis of chronic neuropathic pain based on clinical assessment with a physician, (3) being fluent in English, (4) present consistent medication use for the four weeks leading up to the first baseline visit. Participants were excluded based on the following criteria: (1) having a diagnosis of photosensitive epilepsy, (2) an active ear or eye infection, (3) vision impairments that affect the perception of light, (4) deafness in one or both ears, (5) severe depression.

### 4. Study procedures

Following screening and consenting, participants were randomly assigned to two groups: AVS or Sham. Both groups followed identical protocols, except the AVS group received the active AVS device, while the Sham group received a sham device.

The initial visit included an assessment of all outcome measures, as well as instruction on how to use the tablet to record daily pain and sleep data. At the Week-2 visit, participants repeated their assessments and received instructions on how to use the AVS device at home. Participants were instructed to use their assigned device at least once daily for 8 weeks. Up to 5 additional device uses per day was allowed PRN. After the treatment period of 8 weeks ended, at the Week-10 visit, participants repeated the assessments given at Week-2, were instructed to discontinue use of their assigned device, as well as continue logging daily sleep and pain data. After an additional 4-weeks of non-use, at the Week-14 visit, participants completed their final assessments, and returned the equipment. Outline of the study visits can be seen in Figure 1.

**Figure 1:**
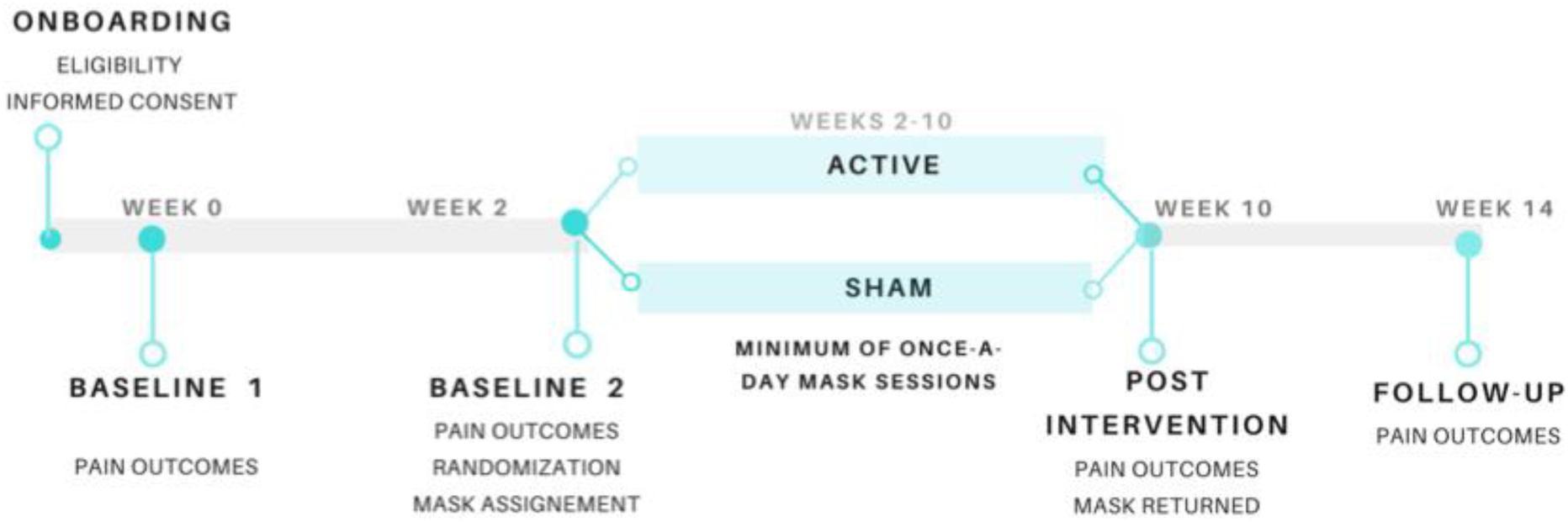
Outline of the protocol and study visits.

### 5. Data collection and outcomes

Data pertaining pain, mood, sleep and quality of life was collected at 4 specific time points: Baseline 1 (week 0), Baseline 2 (week 2), post-treatment (week 10) and follow-up (week 14). In addition, participants were asked to complete daily questionnaires assessing pain, sleep and medication usage throughout the entire study. Assessments were performed in-a clinic or remotely via teleconferencing.

#### A. Primary Outcome Measure

Neuropathic Pain Symptom Inventory (NPSI): Assesses both the quantitative and qualitative properties of neuropathic pain (NP). It includes 12 items, assessing spontaneous pain, brief attacks of pain, provoked pain, and abnormal sensations in the painful area. This is a sensitive tool for measuring changes in neuropathic pain after therapeutic intervention. There are 5 sub-scores (Burning/superficial spontaneous pain, Pressing/deep spontaneous pain, Paroxysmal pain, evoked pain, and Paresthesia/dysesthesia) and a total score ranging from 0-100, with higher scores indicating higher severity [5]. The total score was used as the primary outcome measure.

#### B. Other Outcomes

Secondary and exploratory outcomes were collected in addition to the NPSI to examine changes in common comorbid disorders and other issues associated with NP. Additional outcomes were the Patient Health Questionnaire - 9 (PHQ-9) [28], the Pittsburgh Sleep Quality Index (PSQI) [7], a Sleep Visual Analog Scale (Sleep-VAS) for sleep quality, a Pain Visual Analog Scale (Pain-VAS), the General Anxiety Disorder 7 (GAD-7) for Anxiety [48], and the Patient Global Impression of Change Quality of life Scale (PGIC-QOL) for quality of life [24].

In addition to these outcomes, medications used by patients were recorded and tracked in an electronic log that patients completed daily.

### 6. AVS and Sham Devices

The AVS Device is a goggle style mask used with commercially available headphones. It delivers a patented AVS therapeutic sequence intended to induce deep relaxation and relief from neuropathic pain.

The Sham device is identical in appearance and function to the AVS Device except it only delivers a constant on/off light and tone single sequence with a period of 1.5 seconds. The AVS sequence of the Sham device represents a subset of the range of AVS periods and patterns presented in the full AVS session of the AVS Device. These design features were implemented to produce a convincing sham that would not create a risk of nocebo effects due to the possibility a participant would suspect they received the sham treatment. Thus, the Sham was designed to be a low-dose version of the AVS device and could produce some temporary effects on NP due to temporary relaxation effects and trial related placebo effects. The 4-week non-use period was intended to wash out these temporary effects within the sham arm.

### 7. Data Analysis

All analyses and figures produced using the statistical programming language R [42].

A linear mixed model was fit separately for each outcome. The model had categorical factors of arm (Active/Sham) and visit (up to 6 visits) as well as the interaction between arm by visit. A by-patient intercept was fit to account for correlations across measurements. The main contrasts of interest were comparing the Active to the Sham on change within each outcome after 8 weeks of use and 4 weeks discontinued use in a 2-sided test of superiority. In addition, the effect size for the NPSI was calculated using Cohen’s D for between subject comparisons.

All statistical tests were conducted on 2 analysis populations. The Intent-to-Treat (ITT) population includes any participant that was randomized and completed the baseline assessments. The Per-Protocol (PP) population included any participant that completed the screening, baseline, end-of-treatment, and Week-14 assessments. In addition, they were required to have begun the use of their assigned device and did not have any other major protocol violations.

Device adherence was collected electronically and was calculated as the percentage of device use sessions out of expected device sessions.

#### Calculating the Minimal Clinical Important Difference of the NPSI Total

A Minimal Clinical Important Difference (MCID) value can be used to assess the clinical relevance of a result that is not captured by the statistical comparison of means. The MCID for the NPSI Total is not available in current literature to base an a-priori value on. In the absence of an a-priori MCID value from an external source the FDA recommends the calculation of the MCID using the anchor method [32]. The PGIC-QOL scale is a global patient reported measure of overall improvement collected in this study and was selected as the best available measure to use as the anchor.

To calculate the MCID, we selected the corresponding NPSI change scores within participants that scored into the PGIC-QOL improvement categories of “Minimally Improved” and “Much Improved” at weeks 10 and 14. NPSI change scores from both groups at Week10 and 14 were subset to the scores corresponding to the selected PGIC categories and used in the calculation. To account for the correlation between visits within participants, we used the intercept value of a linear mixed model with a by-patient intercept to produce an estimate of the mean NPSI total change scores. The estimate from the model was a value of 8.29 indicating that an improvement of this magnitude or greater indicates a clinically important change.

#### Medication Change Analyses

Information collected in the daily electronic medication log was used in two analyses. The first analysis looked at changes over the entire study period (Week-2 to Week-14) and the second looked at changes only within the non-use period (Week-10 to Week-14).

Medications that were being used by patients were assigned to the following categories:

1. Anxiolytic and Sedatives
2. Non-Opiate Analgesics
3. Muscle Relaxants
4. Opiate Analgesics
5. Antidepressants
6. Anticonvulsants

The intent of these analyses was to find evidence that improvements on the NPSI in the AVS Arm could have resulted from increased medication use or that lack of improvement for the NPSI within the Sham arm were due to reduced medication use. If a statistical difference between groups within a medication category is found, then the predicted percent change in medication use for both groups will be examined and interpreted. If no between group differences are identified, this suggests that medication changes did not have a differential effect between groups and, thus, no effect on the results of the primary analysis.

For each participant, for each medication, the mean usage within the first 3 days of the trial or the 3 days prior to the Week-10 visit were calculated. The percent change from the 3-day mean was calculated for each subsequent day for each patient for each medication category within both analyses. The data was entered into a linear mixed model which contained a continuous factor for day-mean, a categorical factor for group (Active/Sham), the interaction between day and group, and a by-subject intercept. Statistical difference between Active and Sham will be determined by a statistical separation on the interaction between day-mean and group. Percent change in medication use is the within-group change predicted by the linear model.

## Results

### 1. Participant information

In total, 104 participants were consented for the study. The intention to treat (ITT) group had 64 participants with 31 and 33 in Active and Sham Arms, respectively. Participants at each stage of the study timeline is outlined in Figure 2. Demographic information on the participants in the ITT population can be seen in Table 1. The most common neuropathic pain conditions included in the study were radiculopathy, spinal cord injury, and peripheral neuropathy. Distribution of these between study groups can also be seen in Table 1. Mean length of neuropathic pain for the Active and Sham arms were 4.8 and 8.4 years, respectively.

**Figure 2:**
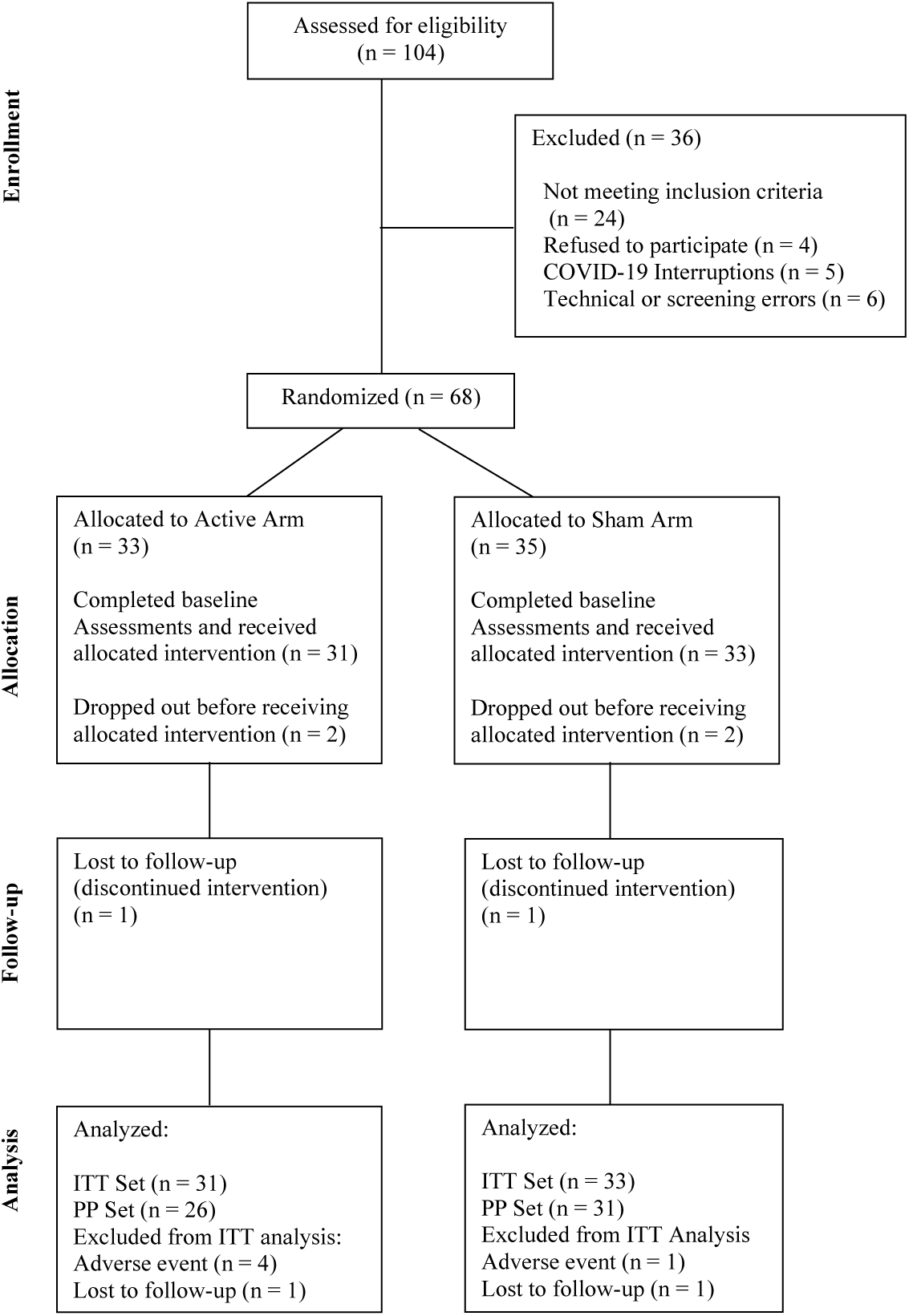
CONSORT flow diagram outlining participants at each stage of the study.

**Table 1:**
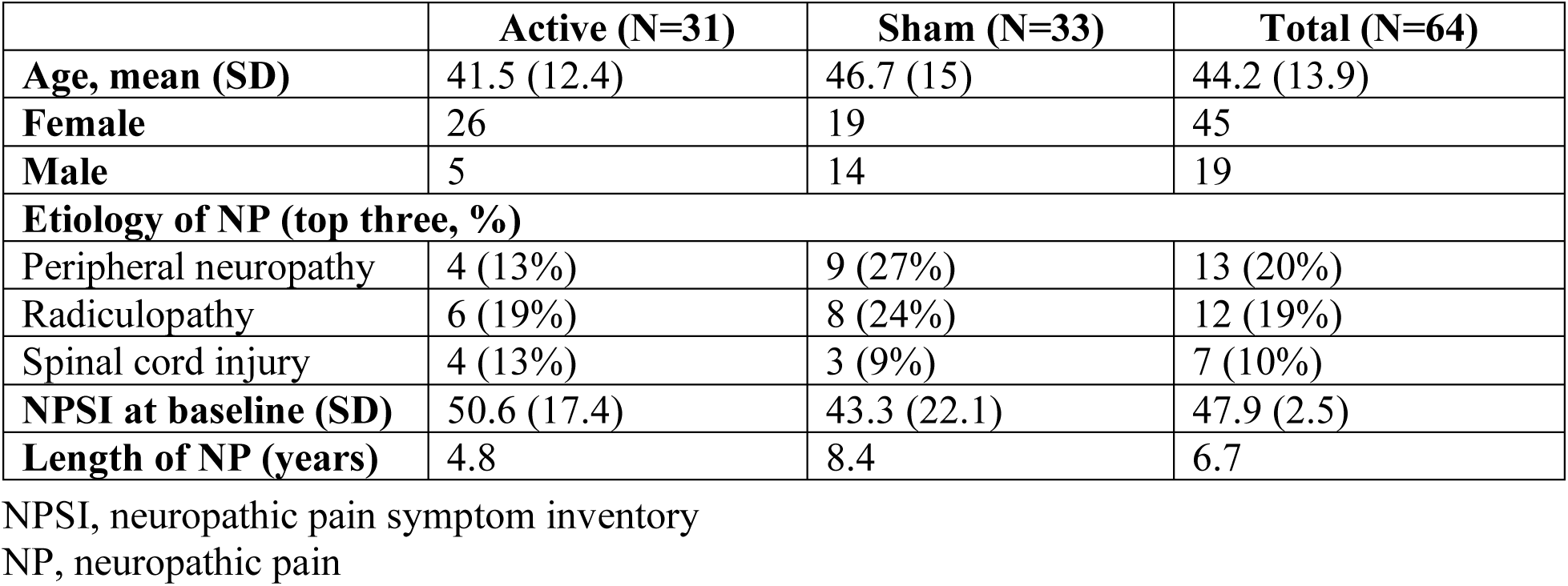
Characteristics of each study group at baseline.

|  | Active (N=31) | Sham (N=33) | Total (N=64) |
| --- | --- | --- | --- |
| **Age, mean (SD)** | 41.5 (12.4) | 46.7 (15) | 44.2 (13.9) |
| **Female** | 26 | 19 | 45 |
| **Male** | 5 | 14 | 19 |
| **Etiology of NP (top three, %)** |  |  |  |
| Peripheral neuropathy | 4 (13%) | 9 (27%) | 13 (20%) |
| Radiculopathy | 6 (19%) | 8 (24%) | 12 (19%) |
| Spinal cord injury | 4 (13%) | 3 (9%) | 7 (10%) |
| **NPSI at baseline (SD)** | 50.6 (17.4) | 43.3 (22.1) | 47.9 (2.5) |
| **Length of NP (years)** | 4.8 | 8.4 | 6.7 |

NPSI, neuropathic pain symptom inventory
NP, neuropathic pain

### 2. Primary Outcome Measure

For the NPSI Total there were statistically significant improvements in the Active arm that were greater than those in the Sham Arm at Week-14, after 8 weeks of treatment and 4 weeks of non-use (ITT: Mean Difference = 10.04, p = 0.01 | PP: Mean Difference = 10.9, p = 0.016) (Figure 3). While both groups showed statistically significant improvements in neuropathic pain at the end of the treatment period (Week-10) compared to baseline (Week-2) (Active Mean Change = 10.02, p <=0.001 | Sham Mean Change = 10.77, p <=0.001), these improvements were not significantly different from each other (Mean Difference = −0.72, p = 0.845). However, after an additional 4 weeks of non-use, at Week-14 only the Active arm maintained this improvement while the Sham arm almost returned to baseline (Active Mean Change = 13.26, p <=0.001 | Sham Mean Change = 3.22, p = 0.214). A summary of NPSI changes within and between study groups is shown in Table 2. The mean change and standard error for the NPSI total for both arms at Week-14 within the ITT population are shown in Figure 4. The effect size for improvement of the Active group over Sham (Cohen’s D for Between Subjects) was calculated to be 0.7, which can be considered a strong effect size.

**Figure 3:**
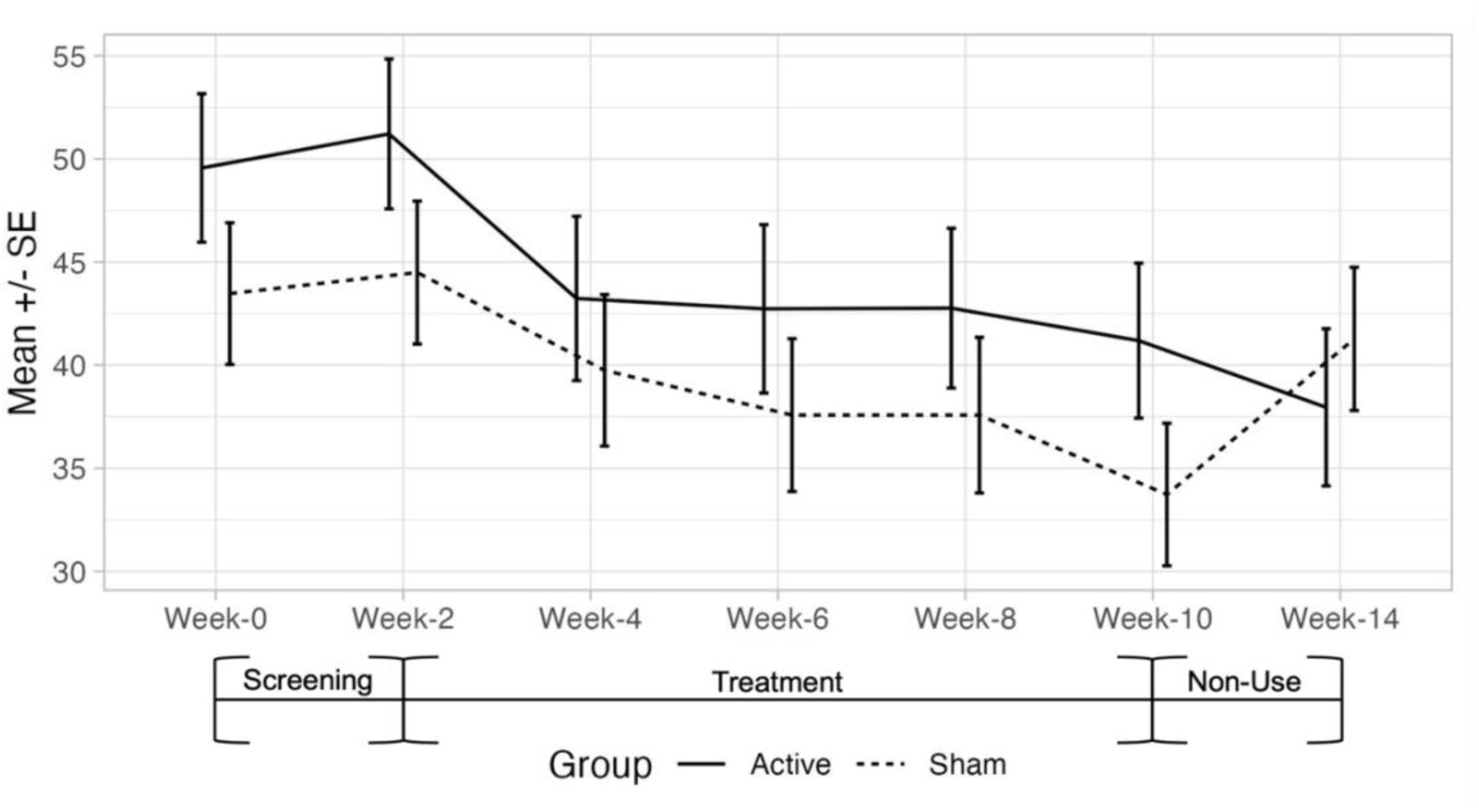
NPSI total for ITT population at each time point during the study. NPSI, neuropathic pain symptom inventory

**Table 2:** NPSI change over time and between groups.

NPSI, neuropathic pain symptom inventory SE, standard error
| Contrast | Mean Change in NPSI | SE | T Ratio | P Value |
| --- | --- | --- | --- | --- |
| Active: Week-2 to Week-10 | 10.024 | 2.809 | 3.569 | <= 0.001 |
| Active: Week-2 to Week-14 | 13.258 | 2.885 | 4.595 | <= 0.001 |
| Sham: Week-2 to Week-10 | 10.765 | 2.559 | 4.207 | <= 0.001 |
| Sham: Week-2 to Week-14 | 3.218 | 2.585 | 1.245 | 0.214 |
| Active(Week10 - Week2) vs Sham(Week10 - Week2) | -0.742 | 3.8 | -0.195 | 0.845 |
| Active(Week14 - Week2) vs Sham(Week14 - Week2) | 10.039 | 3.874 | 2.592 | 0.01 |

**Figure 4:**
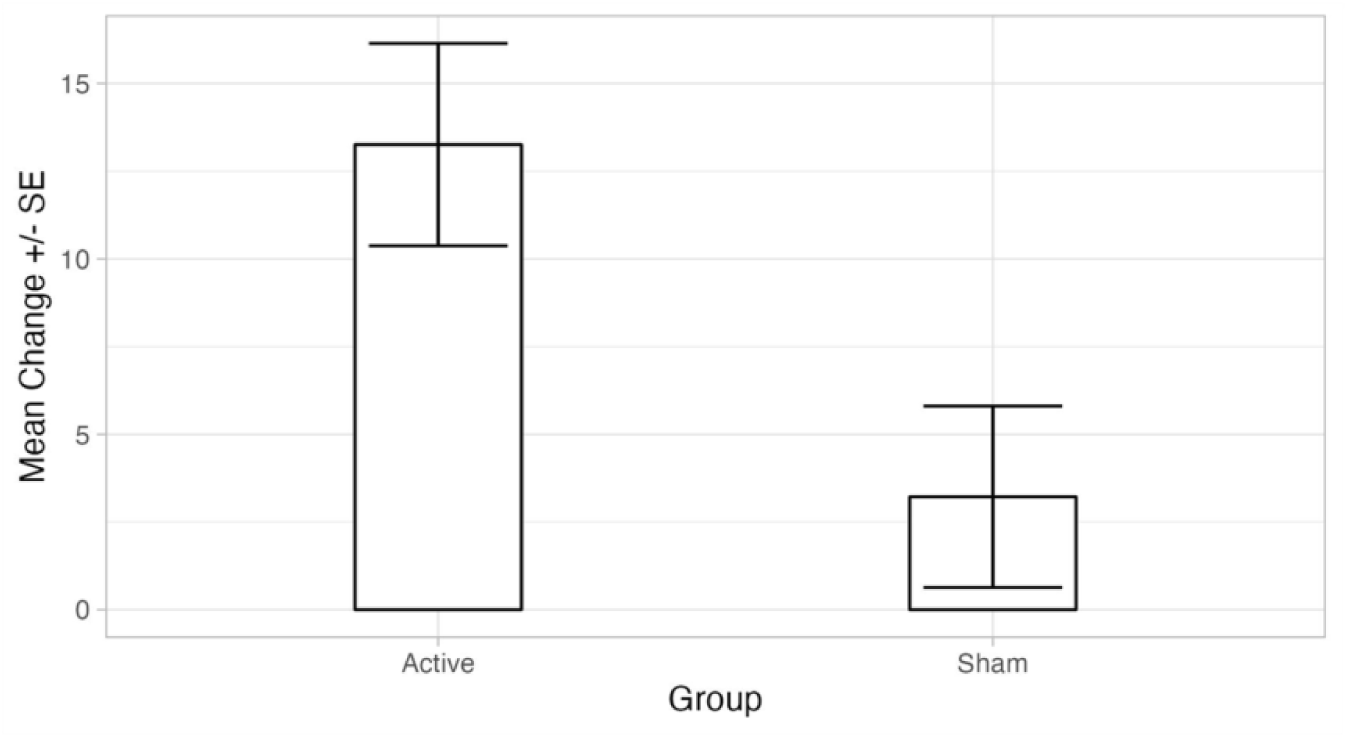
NPSI total: mean change at week-14 by group in ITT population. ITT, intention to treat

Within the NPSI subscales examined, there were statistically significant improvements in the Active arm over sham for the Burning Pain (ITT: Mean Difference = 21.31, p = 0.004; PP: Mean Difference = 22.24, p = 0.008) and Pressing Pain Subscales (ITT: Mean Difference = 13, p = 0.03 | PP: Mean Difference = 11.86, p = 0.1). The mean change and standard error for these subscales for both arms within the ITT population are shown on Figure 5 and Figure 6 respectively. None of the other outcomes examined showed greater improvements in the Active arm over Sham. A summary of all results is presented in Table 3.

**Figure 5:**
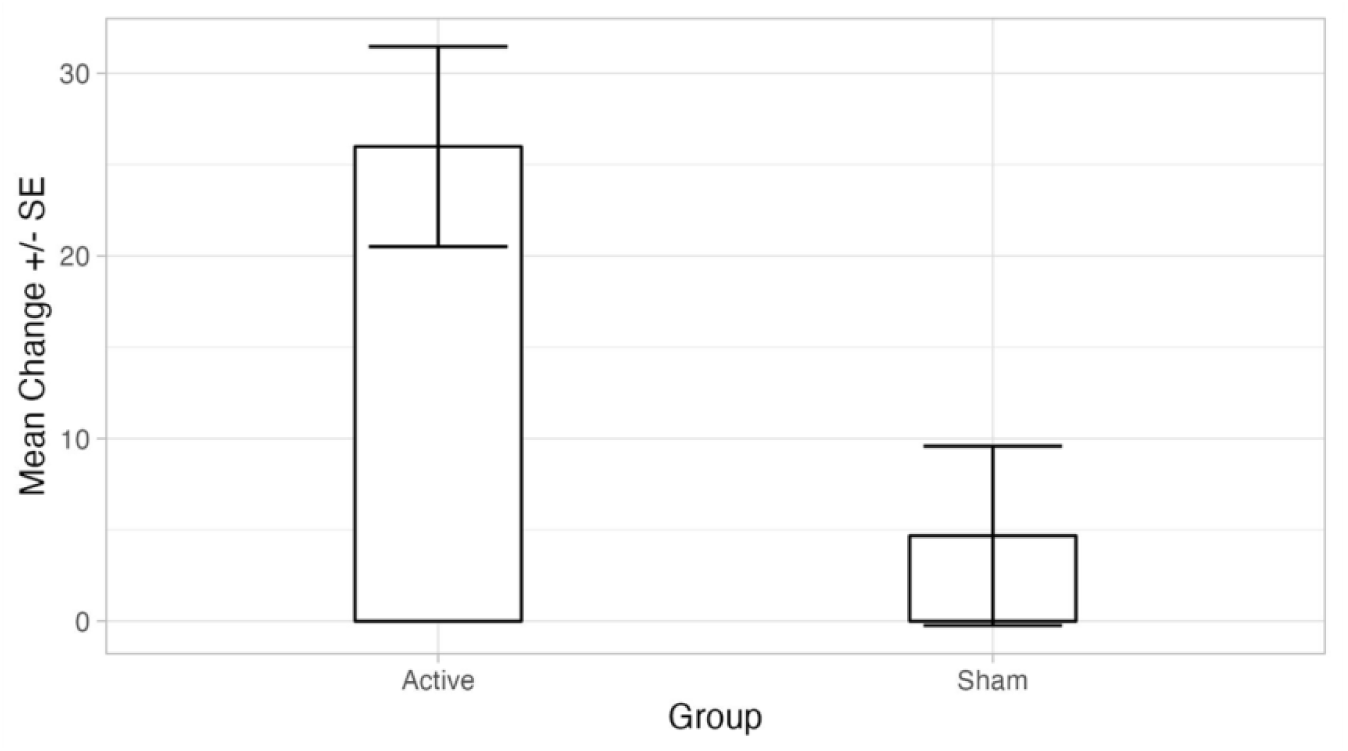
NPSI burning pain subscale: mean change at week-14 by group in ITT population. ITT, intention to treat

**Figure 6:**
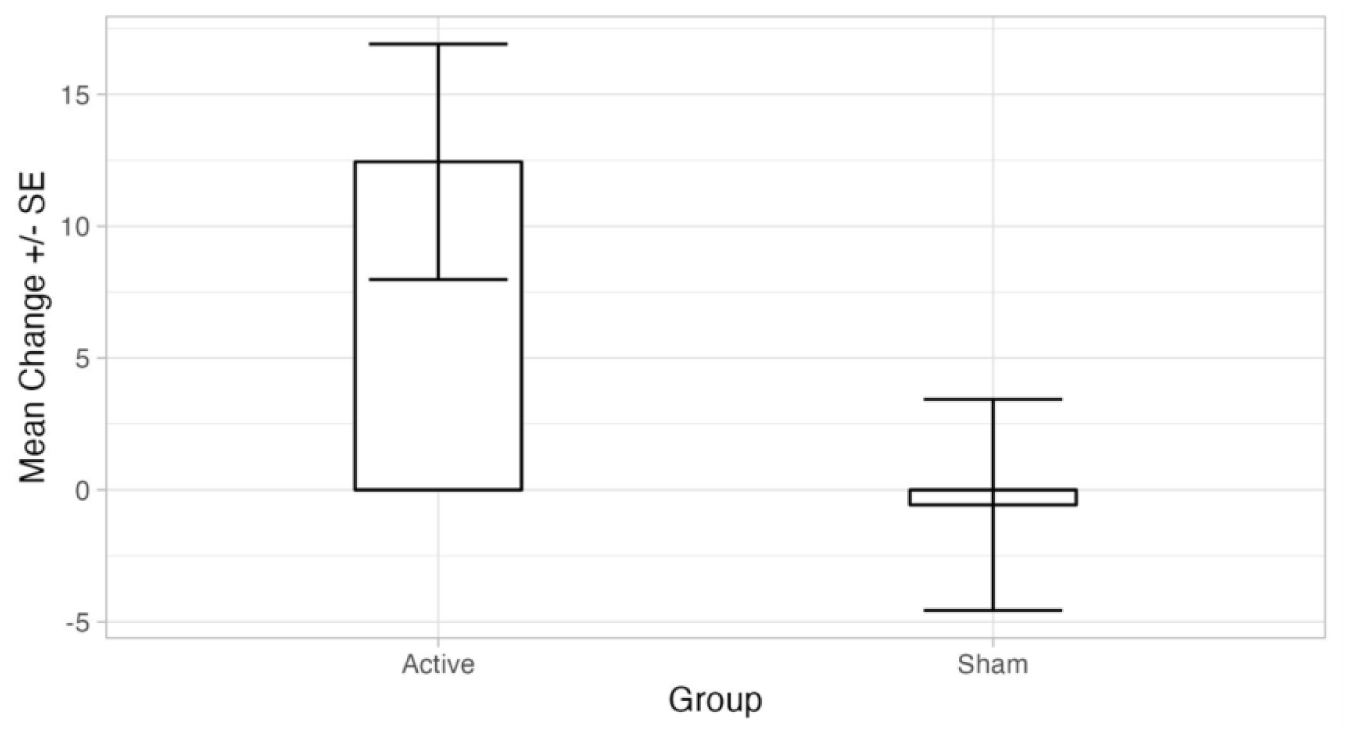
NPSI pressing pain subscale: mean change at week-14 by group in ITT population. ITT, intention to treat

**Table 3:** Breakdown of outcome measures within ITT and PP groups.

| **Outcome** | **Population** | **N**<br>**(Active)** | **N**<br>**(Sham)** | **Mean Difference**<br>**(SE)** | **T**<br>**Ratio** | **P**<br>**Value** |
| --- | --- | --- | --- | --- | --- | --- |
| NPSI Total | ITT | 31 | 33 | 10.039 (3.874) | 2.592 | 0.01 |
|  | PP | 19 | 24 | 10.863 (4.479) | 2.425 | 0.016 |
| NPSI Burning Pain<br>Subscale | ITT | 31 | 33 | 21.312 (7.354) | 2.898 | 0.004 |
|  | PP | 19 | 24 | 22.237 (8.361) | 2.659 | 0.008 |
| NPSI Pressing Pain<br>Subscale | ITT | 31 | 33 | 13.009 (5.995) | 2.17 | 0.03 |
|  | PP | 19 | 24 | 11.864 (7.109) | 1.669 | 0.1 |
| NPSI Evoked Pain<br>Subscale | ITT | 31 | 33 | 4.115 (5.076) | 0.811 | 0.418 |
|  | PP | 19 | 24 | 3.007 (5.905) | 0.509 | 0.611 |
| NPSI Paroxysmal<br>Pain Subscale | ITT | 31 | 33 | 6.121 (6.052) | 1.011 | 0.313 |
|  | PP | 19 | 24 | 8.662 (7.153) | 1.211 | 0.228 |
| NPSI<br>Paresthesia/Dysesthesi<br>a Subscale | ITT | 31 | 33 | 6.327 (5.914) | 1.07 | 0.286 |
|  | PP | 19 | 24 | 8.52 (7.071) | 1.205 | 0.23 |
| GAD-7 Total | ITT | 32 | 39 | -1.305 (0.809) | -1.612 | 0.108 |
|  | PP | 23 | 30 | -1.297 (0.869) | -1.492 | 0.137 |
| PHQ-9 Total | ITT | 32 | 39 | 0.695 (0.847) | 0.82 | 0.413 |
|  | PP | 22 | 29 | 0.702 (0.863) | 0.814 | 0.417 |
| PSQI Total | ITT | 21 | 24 | -0.304 (1.132) | -0.269 | 0.788 |
|  | PP | 17 | 18 | -0.751 (1.265) | -0.594 | 0.553 |
| PGIC-QOL | ITT | 26 | 35 | 0.101 (0.508) | 0.198 | 0.843 |
|  | PP | 25 | 33 | 0.235 (0.53) | 0.444 | 0.659 |

SE, standard error
NPSI, neuropathic pain symptom inventory
GAD-7, General Anxiety Disorder 7 questionnaire
PHQ-9, Patient Health Questionnaire-9
PSQI, Pittsburgh Sleep Quality Index questionnaire
PGIC-QOL, Patient Global Impression of Change Quality of Life scale

### 3. Responder Analysis

A responder in this study is defined as a participant whose improvement on the NPSI Total exceeds the MCID value of 8.29. We calculated the percentage of responders for both Active and Sham at the Week-14 visit within the ITT population (Table 4).

**Table 4:** Percentage of responders in each group, defined as participants whose improvement on NPSI total exceeded the MCID value of 8.29.

|  | **Responders** | **Non-Responders** |
| --- | --- | --- |
| **Active** | 71% (15 of 21) | 29% (6 of 21) |
| **Sham** | 29% (8 of 28) | 71% (20 of 28) |

These values were used in a Chi-Squared (*χ*^2^) of independence. The results of the test indicate that the proportion of responders in the active group is significantly greater than in the Sham group (*χ*^2^ = 7.212, p = 0.007) with the Active group having a response rate of 71% and the Sham group having a response rate of 29%. A test of proportions was used to estimate the upper and lower 95% confidence intervals resulting in a difference in percentage of 43% (95%CI Lower = 13% | 95%CI Upper = 73%). Based on this analysis, the Number Needed to Treat (NNT) with the AVS Device for one person to achieve responder status (mean change >= MCID) is 2.3.

### 4. Device Use and Adherence

Device adherence and average use per day is summarized in Table 5 below. Adherence was defined as using the device at least once per day as described in the study design.

**Table 5:** Mean adherence, defined as using the device at least once per day, and mean sessions per day for each group.

| Group | Mean Adherence (SD) | Mean Sessions per day |
| --- | --- | --- |
| Both (N=64) | 72% (26%) | 1.16 |
| Active (N=31) | 66% (27%) | 1.01 |
| Sham (N=33) | 77% (23%) | 1.29 |

### 5. Change in Medication Usage: Study Duration (Week-2 to Week-14)

The results of the medication log analysis for the study duration indicate significant differences in change of medication usage for the active compared to sham for Anxiolytic and Sedatives (p = 0.009), Muscle Relaxants (p = 0.02) Opiate Analgesics (p = 0.03), Antidepressants (p <= 0.001), and Anticonvulsants (p <= 0.001). See Table 6 below for summary statistics for the Medication Change analysis. A comparison of opiate and anticonvulsant medication use over the study duration for each group can be seen in Figures 7 and 8.

**Table 6:**
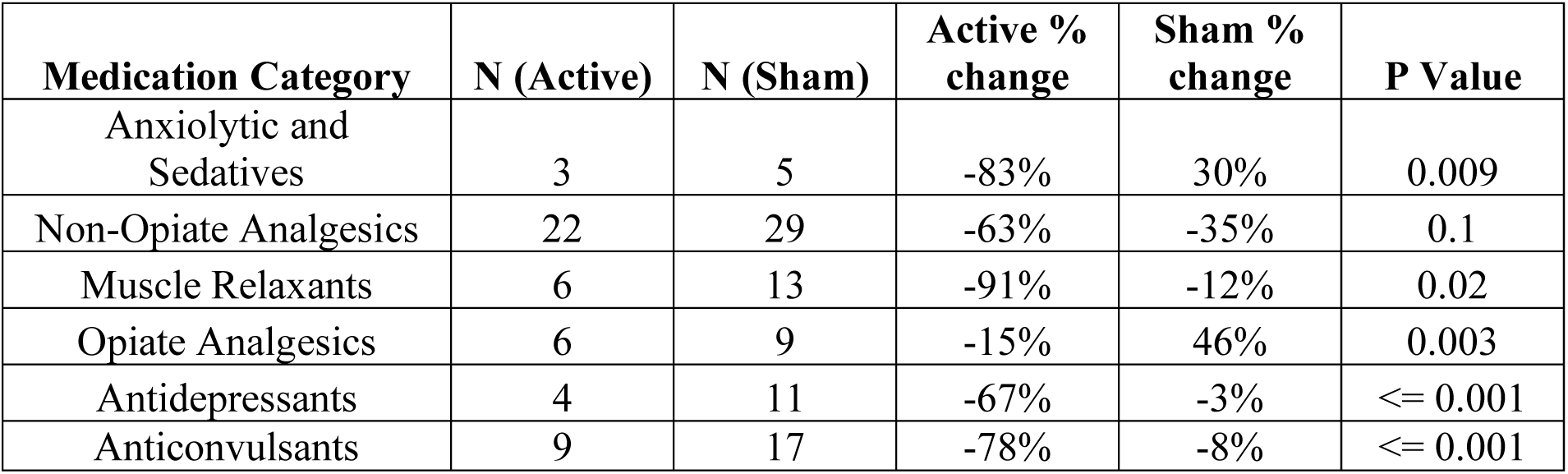
Change in medication use within each group for week-2 to week-14.

| Medication Category | N (Active) | N (Sham) | Active % change | Sham % change | P Value |
| --- | --- | --- | --- | --- | --- |
| Anxiolytic and Sedatives | 3 | 5 | -83% | 30% | 0.009 |
| Non-Opiate Analgesics | 22 | 29 | -63% | -35% | 0.1 |
| Muscle Relaxants | 6 | 13 | -91% | -12% | 0.02 |
| Opiate Analgesics | 6 | 9 | -15% | 46% | 0.003 |
| Antidepressants | 4 | 11 | -67% | -3% | $\leq 0.001$ |
| Anticonvulsants | 9 | 17 | -78% | -8% | $\leq 0.001$ |

**Figure 7:**
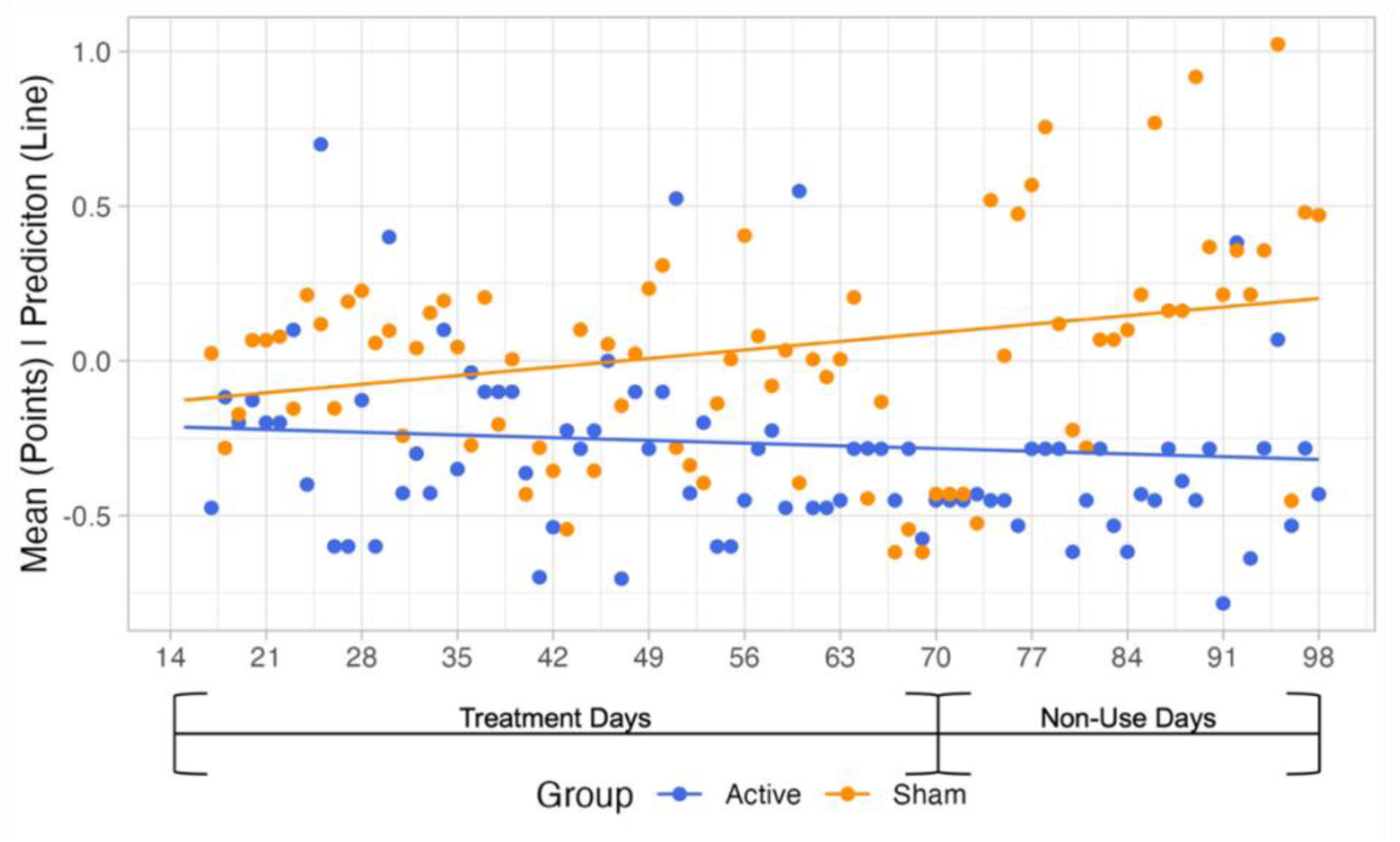
Change in opiate medication usage over the study period.

**Figure 8:**
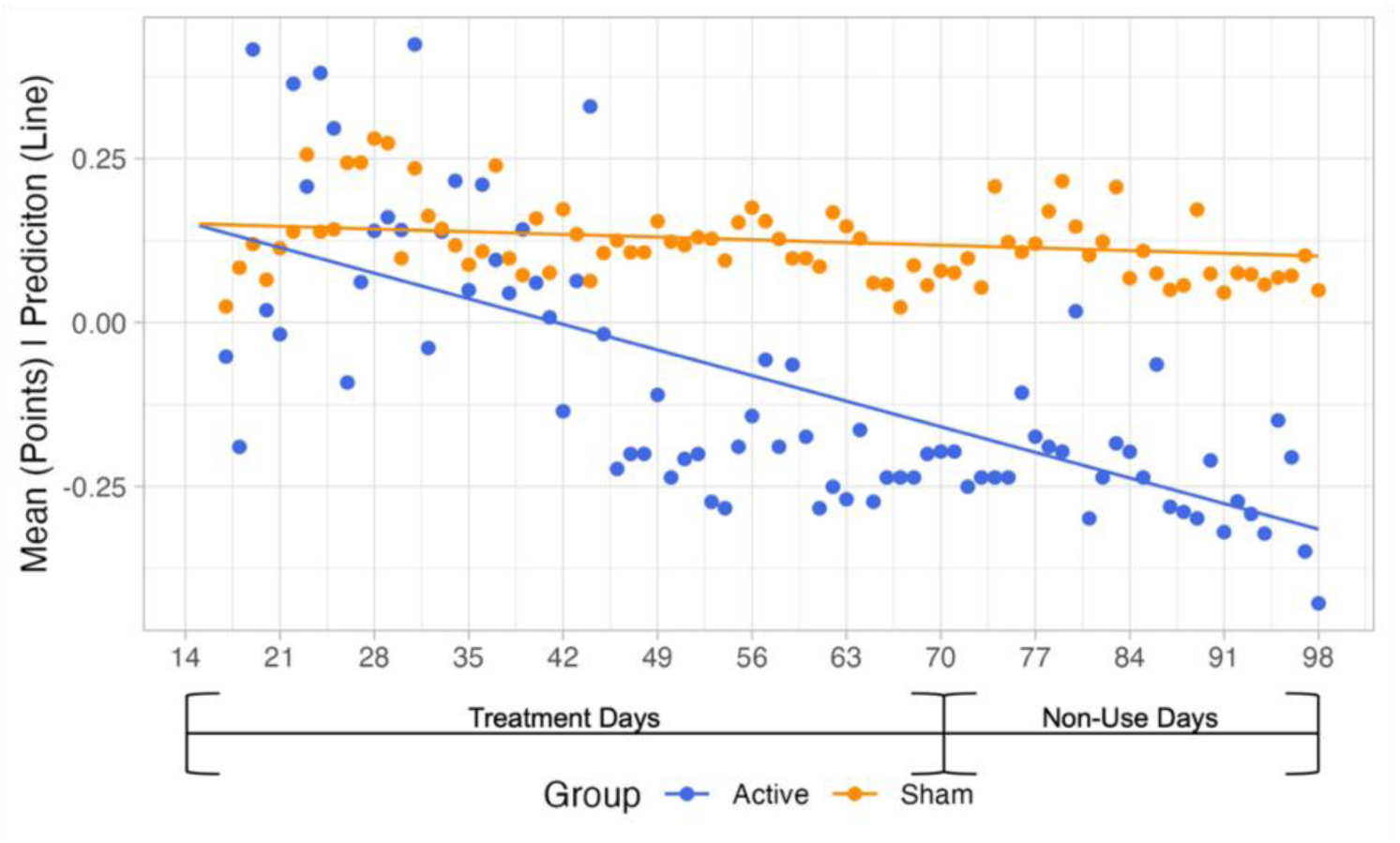
Change in anticonvulsant medication usage over the study period.

### 6. Change in Medication Use: Non-Use Period (Week-10 to Week-14)

The results of the medication log analysis for the non-use period (Week-10 to Week-14) indicate significant changes in the rate of medication use change for the active compared to sham for Anxiolytic and Sedatives (p = 0.035) and Non-Opiate Analgesics (p = 0.002). No other medications showed significant differences in rate of change between Active and Sham. See Table 7 below for summary statistics for the Medication Change analysis.

**Table 7:** Change in medication use within each group from week-10 to week-14.

| **Medication Category** | **N (Active)** | **N (Sham)** | **Active % change** | **Sham % change** | **P Value** |
| --- | --- | --- | --- | --- | --- |
| Anxiolytic and Sedatives | 3 | 5 | -67% | 73% | 0.035 |
| Non-Opiate Analgesics | 22 | 29 | -125% | 22% | 0.002 |
| Muscle Relaxants | 6 | 13 | 0 | -14% | 0.83 |
| Opiate Analgesics | 6 | 9 | 7% | 31% | 0.5 |
| Antidepressants | 4 | 11 | -5% | -6% | 0.92 |
| Anticonvulsants | 9 | 17 | 4% | 10% | 0.66 |

## Discussion

This parallel-arm, blinded, and sham controlled clinical trial evaluated the effectiveness of a novel AVS device compared to a low-dose sham device for reducing the symptoms of neuropathic pain (NP) in 64 participants. Both Active and Sham arms showed statistically significant improvements in NP at the end of the therapeutic period (Week-2 to Week-10), however after an additional 4 weeks of discontinued use, the treatment effect within the Sham arm returned to baseline, while the Active arm maintained their improvement. Efficacy, measured using NPSI total score, was significantly improved in the Active arm compared with the Sham arm, in both ITT and PP populations at Week 14. These results provide strong evidence for the AVS Device in generating and sustaining an improvement in NP, with superiority over Sham at 14 weeks. This improvement is of a clinically significant magnitude, based on the mean change of 10 points shown in the Active arm, which exceeding the estimated anchor based MCID of 8.29. Our responder analysis further supports this, with significantly more responders in the active group (71%) versus sham group (29%) at this MCID. Finally, the Cohen’s D effect size calculation indicates an effect size of 0.7 suggesting a strong effect of the Active device over Sham.

A breakdown of the NPSI subscales revealed that the AVS Device was most effective at improving burning and pressing NP, with significant improvements over Sham for both subtypes at 14 weeks. Based on this pattern of results, it is plausible that the lasting pain relief achieved by the AVS group is primarily attributed to improvements in these manifestations of NP. Both subscales are considered spontaneous ongoing pain and are differentiated by either a superficial (burning) or deep (pressure) sensation. Burning and pressing pain are two of the most common features of NP and are present in up to 70% and 63% of cases, respectively, across a wide range of pathologies, including postherpetic neuralgia, trigeminal neuralgia, radiculopathy, diabetic and HIV neuropathy, amputation, peripheral nerve injury, and stroke [9,10,41,45]. These commonly described pain qualities have a significant impact on quality of life, including sleep, enjoyment of life, depression and anxiety, with all domains more impaired in subjects reporting chronic pain with versus without neuropathic characteristics [3,17]. The pathophysiology behind burning NP involves the firing of unmyelinated C-fibers [22,25]. These C-fibers are a key component of central sensitization, which is a maladaptation of sensory nociceptive pathways which leads to pain hypersensitivity that is not coupled to noxious peripheral stimuli and is considered a form of neuroplasticity [29]. Chronic neuropathic pain has been shown to have a large component of central sensitization, evident in neuroimaging and CSF studies, as well as the use of centrally acting medications as treatment [34,57].

Based on our results, we believe that AVS technology has benefits over conventional therapies for NP in effectiveness, side effect profile, and adherence. Pharmaceutical treatment remains the first line option for both peripheral and central neuropathic pain, with strongest recommendations for anti-epileptics (gabapentin, pregabalin), and antidepressants (serotonin-norepinephrine reuptake inhibitors, tricyclic antidepressants) [1,4]. Despite ongoing research into pharmaceutical options for NP, overall improvement in treatment efficacy has not been observed [12,13]. Clinical effectiveness for these medications is only moderate, based on the number needed to treat (NNT) for 50% pain reduction. In a systematic review and meta-analysis, Finnerup et al. estimated the NNT for Pregabalin to be 7.7, Gabapentin to be 6.3, Tricyclic Antidepressants to be 3.6, and SNRIs to be 6.4 [11]. They suggest that NP is sub-optimally managed, with pharmacologic treatments being effective in <50% of patients. Based on our responder analysis conducted on the NPSI, the NNT for one person to receive clinical benefit after using the AVS Device was calculated to be 2.3. This suggests that fewer people would benefit from treatment with leading pharmaceuticals than from using the AVS Device.

Due to the chronic nature of many patients’ NP, pharmaceutical treatments required for extended periods are often not sustainable due to long-term effects on the kidneys and liver, and burdensome side effects such as dizziness, somnolence, peripheral edema, and dry mouth [2,55]. Adherence remains a challenge, and discontinuation rates for frontline treatments have been reported from 20-60%, both due to insufficient efficacy despite multiple treatments, and unwanted side-effects [1,15,16,35,36,49]. Participants in our study maintained good adherence to using the device at least once per day. In addition, as a non-pharmaceutical product, the AVS device has very minimal side effects, with a total of 3 adverse events deemed to be related to use of the device, and no serious or unanticipated adverse events.

Medication regimens also remain a challenge for treating NP. Patients are more likely to be taking multiple pain medications, including opioids, compared to patients with non-neuropathic chronic pain, and despite this, have less pain relief [38,53]. Additionally, treatment regimens are relatively unstable, with only 30-50% remaining on the same medication over a one-year period [20,38]. Our analysis of changes in medication use showed that overall, participants in the Active arm either decreased or maintained their medication use, while participants in the Sham arm either increased or maintained their medication use. This finding illustrates that the improvements in the Active arm over Sham were obtained even in the presence of decreased medication use in the Active arm and compensatory medication increases the Sham arm. This pattern eliminates the concern that participants changed medication usage in a manner that would explain the results and provides preliminary evidence that the AVS device lowers the need for pain modulating medications including opiates and anticonvulsants.

Participants in the Active arm of the study showed lasting effects on pain reduction even after discontinuation of the AVS device, an effect which was not observed in the Sham arm. One possible explanation for this involves the well described theory of maladaptive neuroplasticity and central sensitization in neuropathic pain [39,58]. AVS technology uses neural entrainment to modulate brain regions associated with pain perception and memory [6]. AVS can target and enhance the amplitude of theta wave activity which plays a role in memory processing and pain modulation [30]. Over time, repeated AVS signals lead to an increased entrainment response suggesting that cortical reorganization has occurred [51]. Given our understanding of AVS, our results support the theory of a central mechanism behind neuropathic pain, with the AVS device potentially reversing some of the maladaptive neuroplasticity associated with NP. The present study was not designed to directly measure neuroplasticity, thus proposals of the mechanism behind the observed effect are speculatory at this time. Future studies that aim to quantify neural changes in response to the device, such as using fMRI, as well as the potential duration of these changes, will be necessary to further shed light on the findings in the present study.

While both study groups had significant improvements in NPSI total score at the end of the treatment period (Week-2 to Week-10), they were not significantly different between groups. This was expected given that the low-dose Sham device provided a combination of distraction, relaxation, and placebo effects. Relaxation techniques have shown to be effective for reducing chronic pain [56], adding a therapeutic value to the Sham device and producing a cumulative, but temporary, mitigation of NP. Developing a true sham device presents challenges when administering a technology such as AVS, where participants are more likely to determine which intervention they received. However, the greater magnitude of improvement seen in the Active arm after a period of non-use suggests that our results cannot be explained by distraction, relaxation, or placebo effects alone. Lack of significant effects of the AVS Device over Sham within the secondary outcomes measuring depression, anxiety, sleep, and quality of life at Week-14 may be due to a partially beneficial Sham device as well. It is likely that any habit that encourages a patient with chronic pain to rest completely for 15-20 minutes multiple times a day will generate positive clinical results. While there was not a statistical difference between the Active and Sham arms for quality of life at Week-14, within the subset of responders in the Active arm, the Cohen’s D effect size (for within subjects) for the PGIC Quality of Life Scale was calculated to be 1.47 which is a very large effect size. This suggests that there was considerable quality of life benefits incurred with the AVS Device.

In conclusion, this randomized controlled trial provides strong evidence for the ability of a novel AVS device to generate durable improvements in overall neuropathic pain intensity that may be driven by improvements in burning pain. In addition, use of the AVS device in this study reduced concurrent use of other commonly used medications for pain. These results highlight the importance for clinicians to accurately characterize patients’ pain, and the need for more precision medicine targets for specific pain subtypes. Overall, the excellent device usage adherence and low rate of device related adverse events are supportive of the AVS device as a safe, easy to use, and effective treatment for reducing the severity of neuropathic pain.

## Data Availability

Data are available from the authors upon reasonable request. Please contact for data inquiries.

## Acknowledgements

Funding for the study was provided by Sana Health, Inc. At the time of submission, Richard Hanbury holds the title of CEO of Sana Health, Inc, and Jeffrey Bower holds the title of VP of Analytics at Sana Health, Inc.

The other authors have no conflicts of interest to declare.

Data are available from the authors upon reasonable request. Please contact for data inquiries.

